# Psychopharmacoepidemiological trends in the Portuguese territory during the pandemic and post-pandemic periods (2019–2022)

**DOI:** 10.1101/2024.05.26.24307948

**Authors:** David Belchior, Luís Madeira, Rui Henriques

## Abstract

The increasing prevalence of mental health disorders has been matched with growing psychotropic drug consumption rates around the world. Assessing psychopharmacoepidemiological trends and their determinants is essential to guide medical care delivery and public health policies. However, nation-wide studies on up-to-date consumption patterns are scarce and generally disregard important pharmacological, medical, sociodemographic, and economic covariates. Previous studies on the Portuguese case, a case known for critically high consumption rates of benzodiazepines and antidepressants, are limited to the COVID-19 pre-pandemic period. This study uses the full (electronic) dispensation registry of antidepressants, benzodiazepines and zolpidem, antipsychotics and mood stabilisers in Portugal during the years of 2019 to 2022 with the goal of identifying relevant prescription and consumption patterns prior to, during and after the COVID-19 pandemic. Our findings show a consumption growth trend in antidepressants (7.41% yearly DIDs, *P* = 0.0215) accelerated since 2020, confirmed by the growing number of users (over 15% of the population), overtaking benzodiazepines and zolpidem as the class with most active users. The total annual expenditure has increased 14M€ between 2020 and 2022 (nearly 2M€ in public copayment), notwithstanding price drops in diverse antipsychotic drugs.

## 1 INTRODUCTION

Mental health conditions currently rank among the most serious non-communicable diseases by disability-adjusted life years per year [16], with unprecedented prevalence growth trends [9]. In particular, Portugal is the fourth country in the European Union with highest estimated risk of depression in the adult population (with 61% of the population at risk in 2022), and the second country across OECD members with highest estimated consumption of antidepressants [31]. High prescription rates in the Portuguese case are also observed for other psychopharmacological classes, in particular, benzodiazepines. However, there is no comprehensive assessment of the COVID-19 pandemic trends in consumption and expenditures, as well as their underlying covariates.

This study aims at profiling nation-wide psychopharmacoepidemiological trends in the Portuguese territory as a function of relevant covariates, including the geographic location, patient’s demographic profile, pharmacological properties, and prescriber’s specialty. To this end, we use the complete nation-wide dispensation registry on antidepressants, benzodiazepines and zolpidem, antipsychotics, mood stabilisers and pregabalin on the Portuguese Ministry of Health’s Prescrição Eletrónica de Medicamentos (PEM) system from 2019 to 2022. This offers the opportunity to assess pharmacological, medical, demographic and geographic determinants during and after the pandemic period, as a way to guide the design and implementation of public policy actions.

## 2 RELATED WORK

In recent years, a variety of cross-generational retrospective studies on the analysis of psychotropic drug consumption trends have been undertaken. In the case of Portugal, a study dedicated to the pre-pandemic 2016–2019 period [27] revealed an increase of 29.6% and 34.7% in the consumption (in DIDs^1^) of antipsychotics and antidepressants, respectively, with an increase of 37 M€ in expenditures, over 20 M€ of which coming from state-funded copayment. As for the ratio of prescriptions by medical specialty, 64% of psychotropic drug prescriptions were undertaken by general practitioners (GPs), while only 21% were undertaken by neurological and psychiatric specialties. We aim to directly continue this study by extending the analysed data towards the 2020–2022 period, encompassing the effects of the COVID-19 pandemic on the Portuguese society.

Other countries have reported generalised increased consumption of psychotropic drugs during and after the COVID-19 pandemic, namely Croatia [39], France [3, 22], Spain [11] and Brazil [7].

Amongst the varying approaches to quantify drug consumption incidence, the DID is frequently suggested as a reference proxy for incidence [5, 27], providing a uniform, drug-independent perspective on a population’s consumption patterns. Notwithstanding its relevance for precise nation-wide assessments, most studies are constrained by observability limitations such as the restriction to subpopulations [23, 38], the reliance on self-reported data [5], or outpatient prescriptions [27, 38].

## 3 METHODS

### Data collection

This work undertakes a nation-wide retrospective study centred on the complete dispensation registry acquired by the PEM platform between 2019-01-01 and 2022-12-31 on psychotropic drugs with Infarmed’s approval for commercial use, containing data related to all dispensations of Portuguese citizens aged 18 or older, excluding those who were never dispensed before the moment of data collection. The full list of considered drugs, divided by class and subclass, including their ATC code (if existent) and DDD (defined by route of administration), as well as their various commercialised packages, can be found in the appendix.

The drug categorisation is aligned with ATC classification, with some exceptions. Benzodiazepines and zolpidem are grouped into a single category, regardless of their additional classification as anxiolytics or sedative/hypnotics; this includes the combination of chlordiazepoxide with clinidium bromide, an antispasmoid, with the considered DDD matching chlordiazepoxide’s as a standalone drug. Pirlindole has no ATC code as of yet, and thus has an assigned DDD value based on previous studies with the same active ingredient. As valproic acid and lamotrigine (classified as antiepileptics) are also administered as mood stabilisers, they are included in this class. Pregabalin is also analysed, albeit individually.

In order to standardise demographic deviations across the Portuguese territory, data regarding Portugal’s administrative divisions (municipalities, districts and NUTS) and their population is retrieved from INE, Portugal’s statistics institute [17].

### Data processing

The data warehousing process, necessary for a quick retrieval of dispensation patterns, entails a processing stage, where the raw data is cleaned and harmonised between the different data sources (prescriptions, drugs, packaging, medical specialities, and demographics) before their consolidation. The system developed for the extraction, transformation and loading of the data, as well as its agglomeration into the metrics and the resulting graphs, is divided into three major layers: a PostgreSQL relational database management system (RDBMS); a Django back-end responsible for processing the raw data, aggregating it and exposing it with dedicated API endpoints by communicating with the database; a Python front-end for creation of data requests and the conversion of their results into dataframes, which are then plotted using Matplotlib.

The dispensation data includes only 7 age groups, spanning 10 years each, with the exception of the first (18-29) and last (80 or older) groups; this contrasts with the yearly (estimated) population data retrieved from INE, which is divided in 18 age groups, spanning 5 years each up to the final group, which includes people aged 85 or older, thus requiring modifications to align both sources. First, the 15-19 age group is divided into the 15-17 and the 18-19 groups. Considering yearly birth statistics, this division is made assuming a yearly uniform population distribution, meaning the 18-19 group contains 40% of the initial count (rounded up), which is added to the 20-24 and 25-29 groups, creating the 18-29 group. Second, the 80-84 and 85 or older groups are merged in a single 80+ age group. Finally, the remaining age groups are presented in 10-year segments.

Each individual dispensation act includes the dispensed drug’s information, the dispensation’s full price and governmental copayment, the prescriber’s speciality, a timestamp (year and week), and the patient’s (anonymous) ID, age group and gender.

Each dispensation act’s location is determined by the pharmacy where it happened or, when unavailable, by the corresponding prescription act’s location (e.g., a hospital or healthcare centre).

Data from zolpidem, cariprazine and clozapine only features in the supplied data since 2020, despite their previous approval by Infarmed. In this context, to provide a more statistically accurate estimate on dispensed DIDs and overall costs on their respective classes, the dispensation of these specific drugs is linearly extrapolated for the year of 2019.

Region-specific statistics related to dispensed DIDs are standardised by the national distribution of demographic sections using the *k* coefficient,

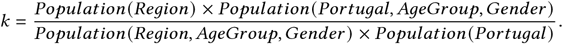

#### 3.1 Statistical analysis

Trends were tested at a 5% significance threshold for the regression coefficient under a two-sided hypothesis on the regression coefficient with a null hypothesis that the mean year-on-year rate of change is zero, using the Wald test with a t-distribution of the test statistic.

## 4 RESULTS

The most relevant results can be found in Tables 1–4 and Figures 1–6.

**Table 1.** Evolution of dispensed DIDs in benzodiazepines and zolpidem between 2019 and 2022.

| Gender | Age group | 2019 | 2020 | 2021 | 2022 | Mean change per year,<br>fraction of baseline | P | 95% confidence interval | r<sup>2</sup> |
| --- | --- | --- | --- | --- | --- | --- | --- | --- | --- |
| F | 18-29 | 0.6370 | 0.7056 | 0.7841 | 0.8145 | 0.0959 | 0.0140 | [0.0466, 0.1452] | 0.9722 |
|  | 30-39 | 1.8861 | 2.0130 | 1.9954 | 1.9352 | 0.0069 | 0.7118 | [-0.0625, 0.0763] | 0.0830 |
|  | 40-49 | 5.5923 | 5.9108 | 5.9941 | 5.8082 | 0.0131 | 0.4563 | [-0.0483, 0.0745] | 0.2956 |
|  | 50-59 | 9.3648 | 9.7367 | 9.9477 | 9.8139 | 0.0166 | 0.1942 | [-0.0206, 0.0538] | 0.6492 |
|  | 60-69 | 11.6877 | 12.1447 | 12.5675 | 12.5468 | 0.0257 | 0.0656 | [-0.0041, 0.0555] | 0.8731 |
|  | 70-79 | 11.2125 | 11.4917 | 11.9762 | 12.0590 | 0.0270 | 0.0294 | [0.0067, 0.0473] | 0.9422 |
|  | 80+ | 9.4550 | 9.5568 | 9.7701 | 9.6668 | 0.0090 | 0.1959 | [-0.0112, 0.0292] | 0.6466 |
|  | **Total** | 49.8355 | 51.5593 | 53.0351 | 52.6444 | 0.0199 | 0.1072 | [-0.0106, 0.0504] | 0.7970 |
| M | 18-29 | 0.5263 | 0.5738 | 0.6132 | 0.6442 | 0.0747 | 0.0044 | [0.0534, 0.0960] | 0.9913 |
|  | 30-39 | 1.3444 | 1.4144 | 1.4260 | 1.3996 | 0.0132 | 0.3663 | [-0.0358, 0.0622] | 0.4016 |
|  | 40-49 | 3.0552 | 3.1927 | 3.2644 | 3.2277 | 0.0193 | 0.1673 | [-0.0197, 0.0583] | 0.6935 |
|  | 50-59 | 4.2531 | 4.4622 | 4.6425 | 4.6567 | 0.0327 | 0.0499 | [0.0000, 0.0654] | 0.9027 |
|  | 60-69 | 5.0707 | 5.2153 | 5.3490 | 5.3999 | 0.0221 | 0.0190 | [0.0088, 0.0354] | 0.9625 |
|  | 70-79 | 4.7381 | 4.8671 | 5.0427 | 5.0771 | 0.0252 | 0.0270 | [0.0070, 0.0434] | 0.9468 |
|  | 80+ | 3.0731 | 3.1077 | 3.1548 | 3.1443 | 0.0085 | 0.0954 | [-0.0037, 0.0207] | 0.8183 |
|  | **Total** | 22.0608 | 22.8332 | 23.4925 | 23.5496 | 0.0232 | 0.0493 | [0.0001, 0.0463] | 0.9038 |
| **Total** |  | 71.8963 | 74.3924 | 76.5276 | 76.1940 | 0.0209 | 0.0860 | [-0.0073, 0.0491] | 0.8355 |

**Table 2.**
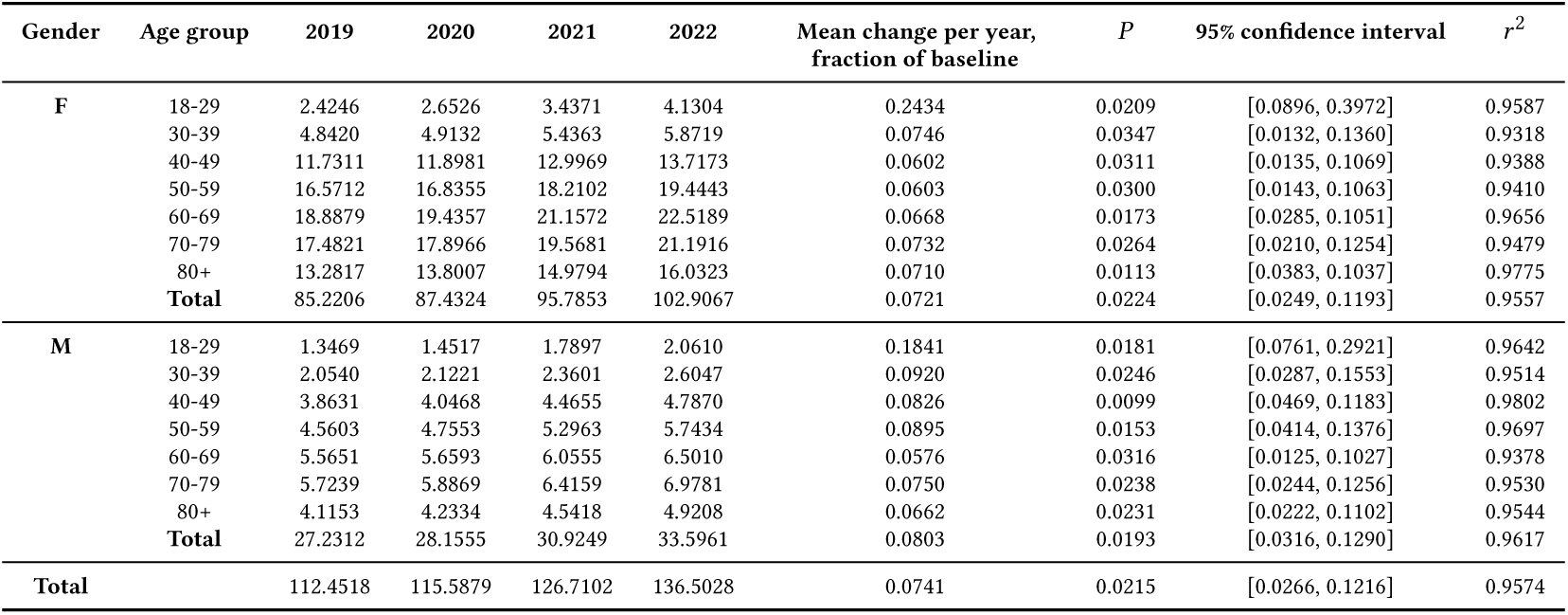
Evolution of dispensed DIDs in antidepressants between 2019 and 2022.

**Table 3.**
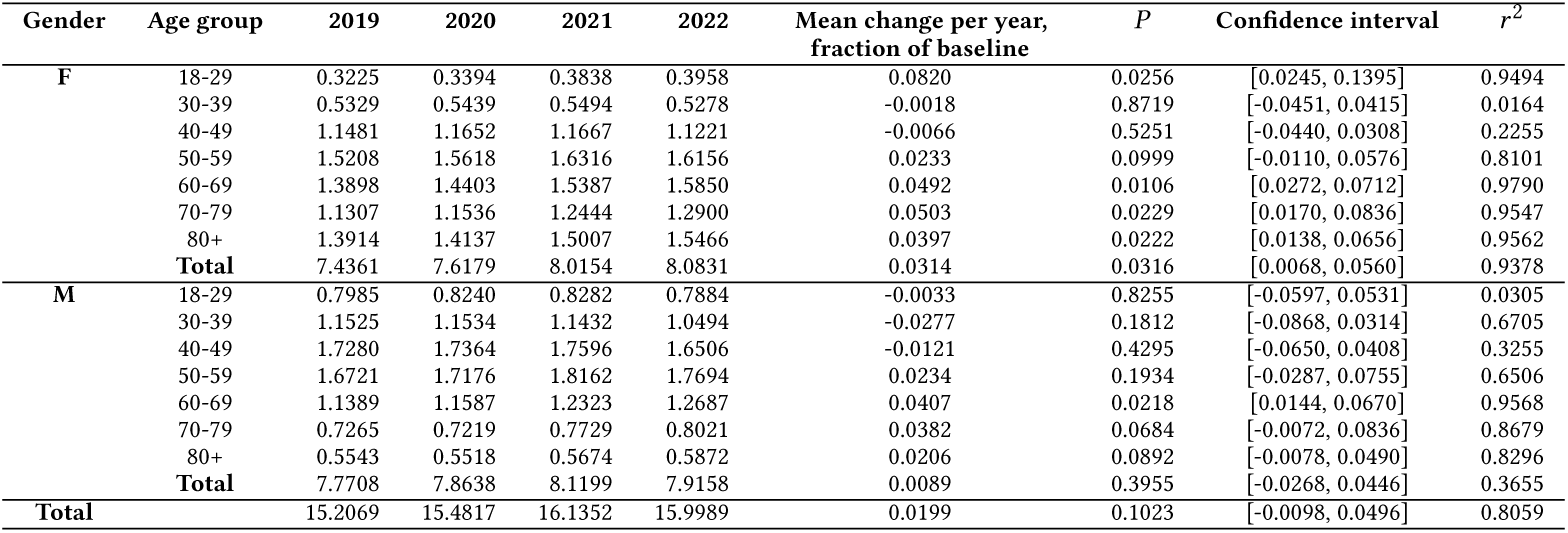
Evolution of dispensed DIDs in antipsychotics between 2019 and 2022.

**Table 4.** Evolution of dispensed DIDs in mood stabilisers between 2020 and 2022.

| Gender | Age group | 2020 | 2021 | 2022 | Mean change per year,<br>fraction of baseline | P | 95% confidence interval | r<sup>2</sup> |
| --- | --- | --- | --- | --- | --- | --- | --- | --- |
| F | 18-29 | 0.1456 | 0.1554 | 0.1667 | 0.0724 | 0.0254 | [0.0356, 0.1092] | 0.9984 |
|  | 30-39 | 0.2023 | 0.2035 | 0.2102 | 0.0197 | 0.2450 | [-0.0818, 0.1212] | 0.8591 |
|  | 40-49 | 0.3778 | 0.3718 | 0.3758 | -0.0027 | 0.7864 | [-0.0996, 0.0942] | 0.1084 |
|  | 50-59 | 0.4344 | 0.4525 | 0.4624 | 0.0322 | 0.1086 | [-0.0383, 0.1027] | 0.9712 |
|  | 60-69 | 0.3577 | 0.3710 | 0.3822 | 0.0342 | 0.0315 | [0.0127, 0.0557] | 0.9975 |
|  | 70-79 | 0.1860 | 0.1968 | 0.2064 | 0.0551 | 0.0225 | [0.0304, 0.0798] | 0.9988 |
|  | 80+ | 0.0792 | 0.0787 | 0.0787 | -0.0030 | 0.3778 | [-0.0290, 0.0230] | 0.6872 |
|  | Total | 1.7829 | 1.8297 | 1.8823 | 0.0279 | 0.0218 | [0.0158, 0.0400] | 0.9988 |
| M | 18-29 | 0.2031 | 0.2129 | 0.2103 | 0.0177 | 0.4991 | [-0.2060, 0.2414] | 0.5014 |
|  | 30-39 | 0.2361 | 0.2365 | 0.2322 | -0.0083 | 0.3824 | [-0.0809, 0.0643] | 0.6806 |
|  | 40-49 | 0.3323 | 0.3370 | 0.3337 | 0.0022 | 0.8090 | [-0.0871, 0.0915] | 0.0873 |
|  | 50-59 | 0.3216 | 0.3348 | 0.3390 | 0.0271 | 0.1859 | [-0.0762, 0.1304] | 0.9171 |
|  | 60-69 | 0.2352 | 0.2409 | 0.2468 | 0.0248 | 0.0056 | [0.0220, 0.0276] | 0.9999 |
|  | 70-79 | 0.1373 | 0.1416 | 0.1444 | 0.0258 | 0.0820 | [-0.0167, 0.0683] | 0.9835 |
|  | 80+ | 0.0481 | 0.0491 | 0.0500 | 0.0194 | 0.0075 | [0.0165, 0.0223] | 0.9999 |
|  | Total | 1.5136 | 1.5528 | 1.5564 | 0.0141 | 0.2860 | [-0.0723, 0.1005] | 0.8114 |
| Total |  | 3.2965 | 3.3825 | 3.4387 | 0.0216 | 0.0766 | [-0.0115, 0.0547] | 0.9856 |

**Fig. 1.**
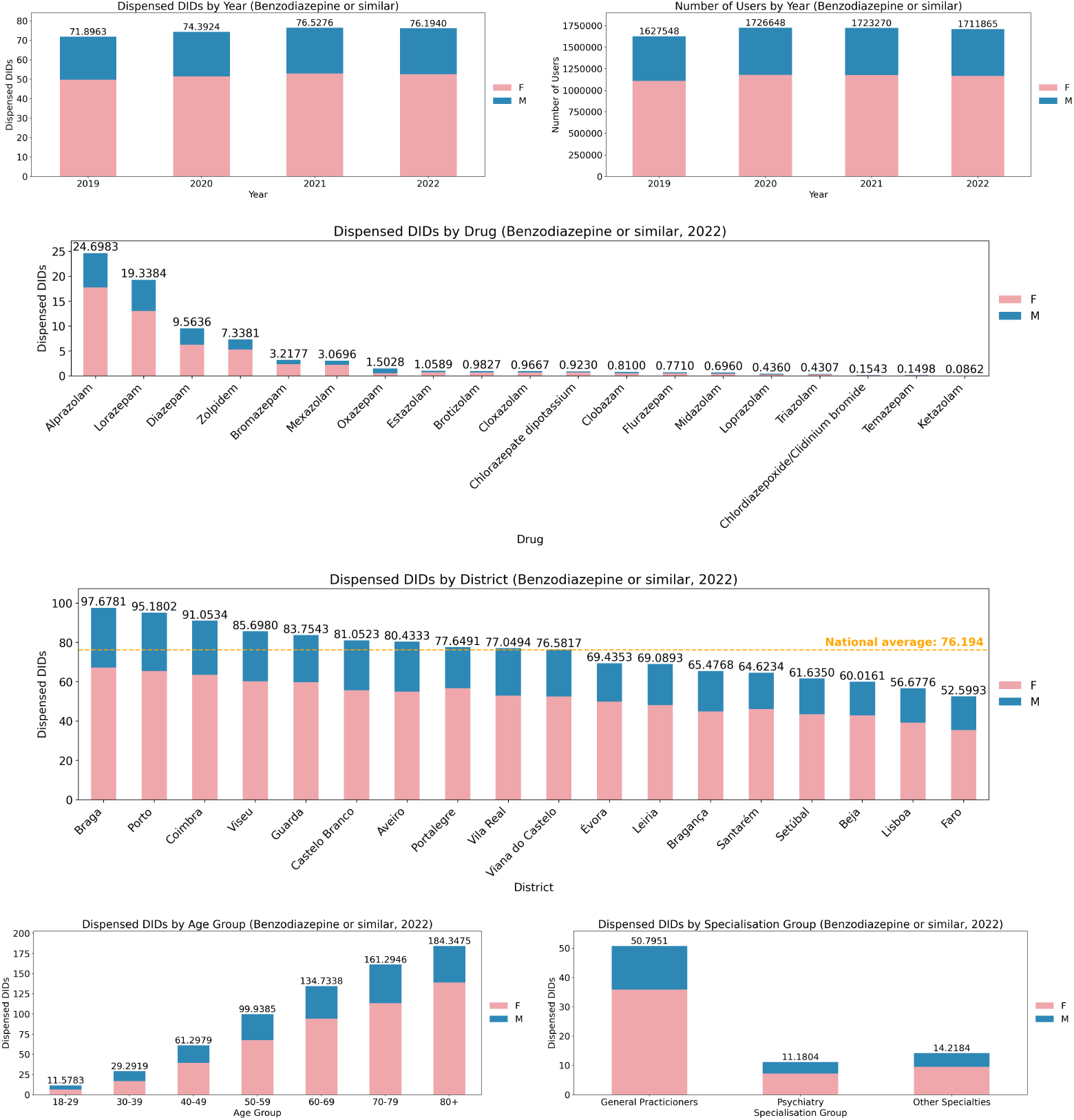
Benzodiazepine and zolpidem dispensation statistics (DIDs and user count) across districts, demo-graphic sectors and medical specialties (gender is presented using stacked representations).

**Fig. 2.**
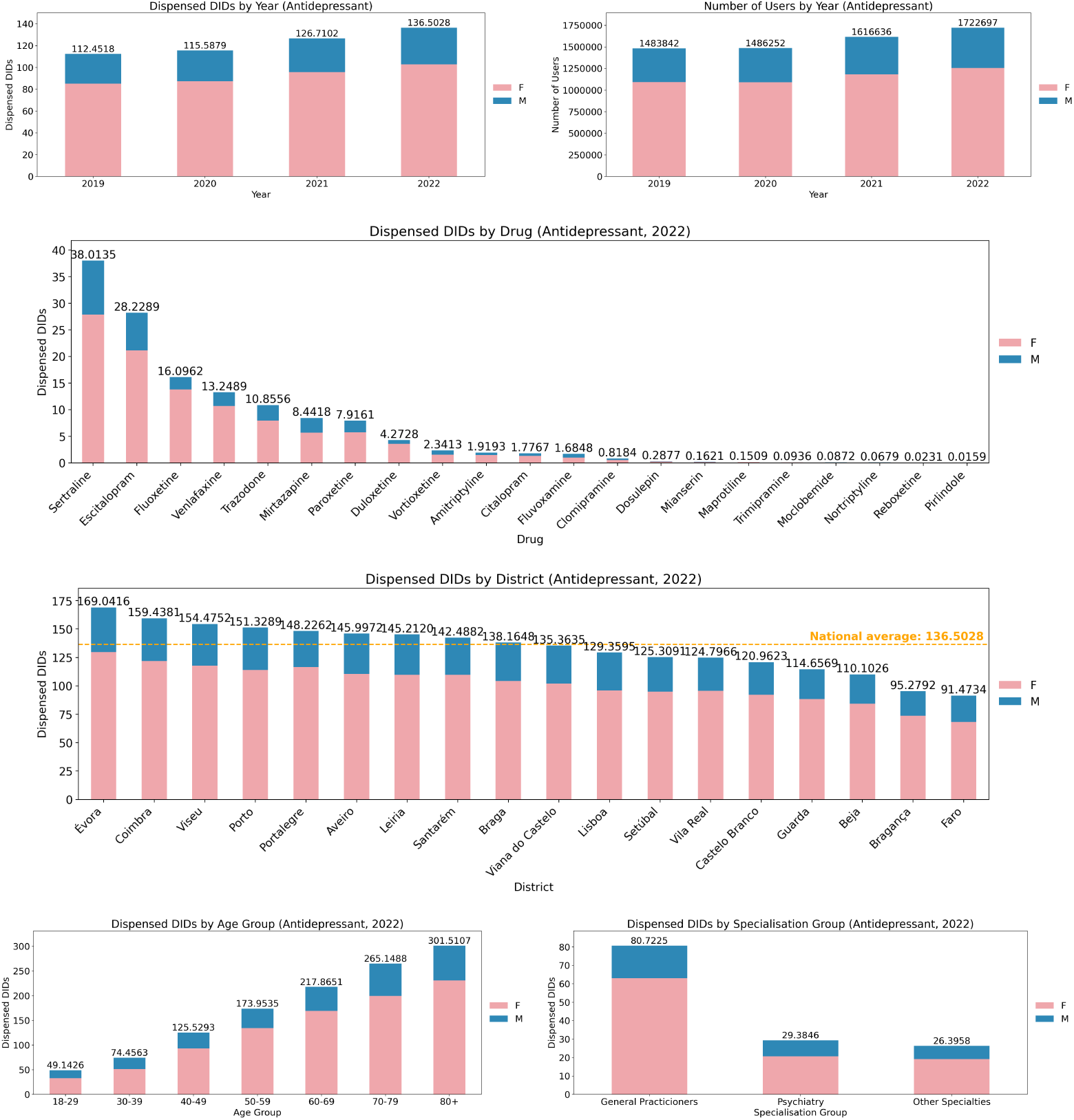
Antidepressant dispensation statistics (DIDs and user count) across districts, demographic sectors and medical specialties (gender is presented using stacked representations).

**Fig. 3.**
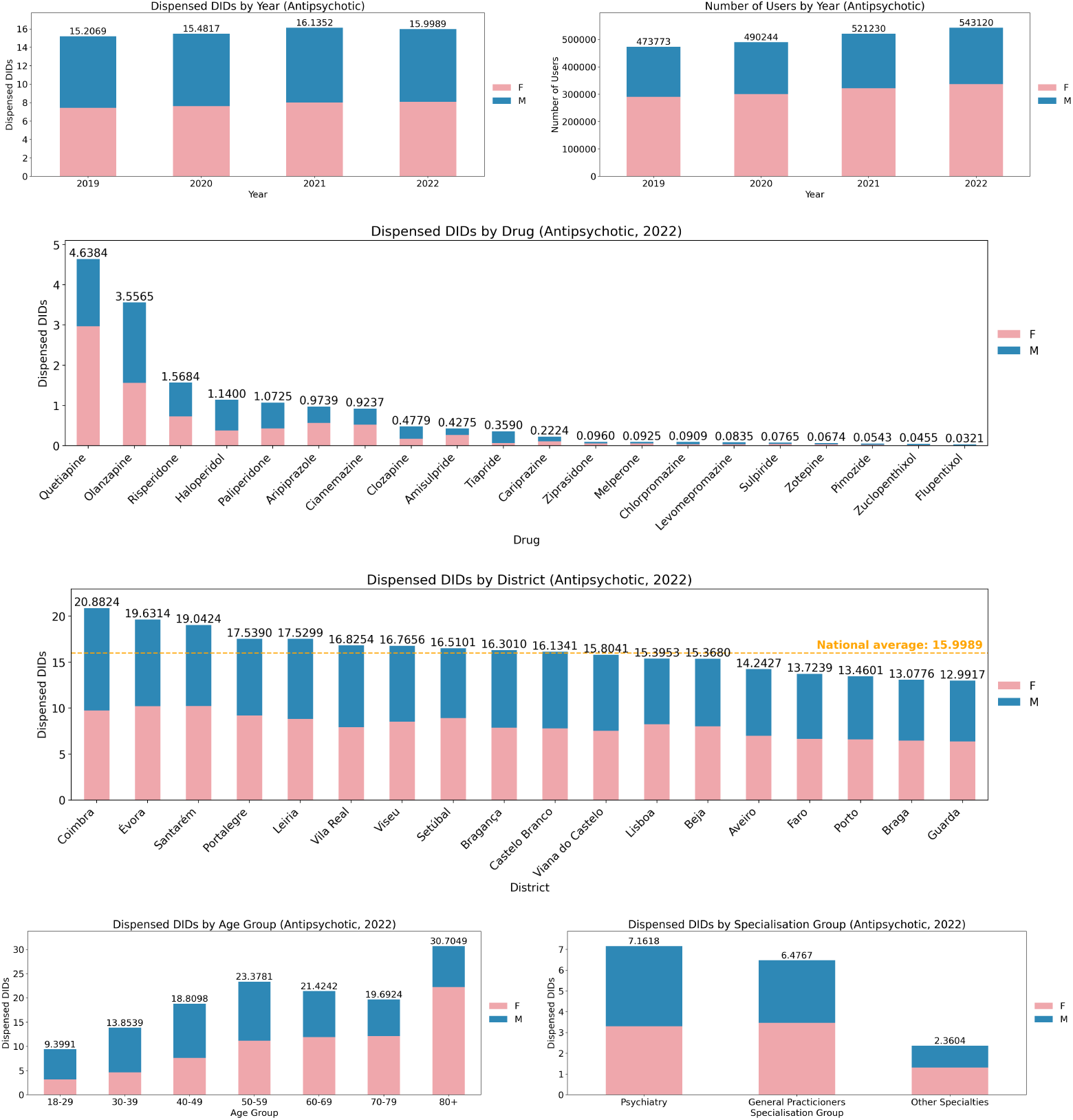
Antipsychotic dispensation statistics (DIDs and user count) across districts, demographic sectors and medical specialties (gender is presented using stacked representations).

**Fig. 4.**
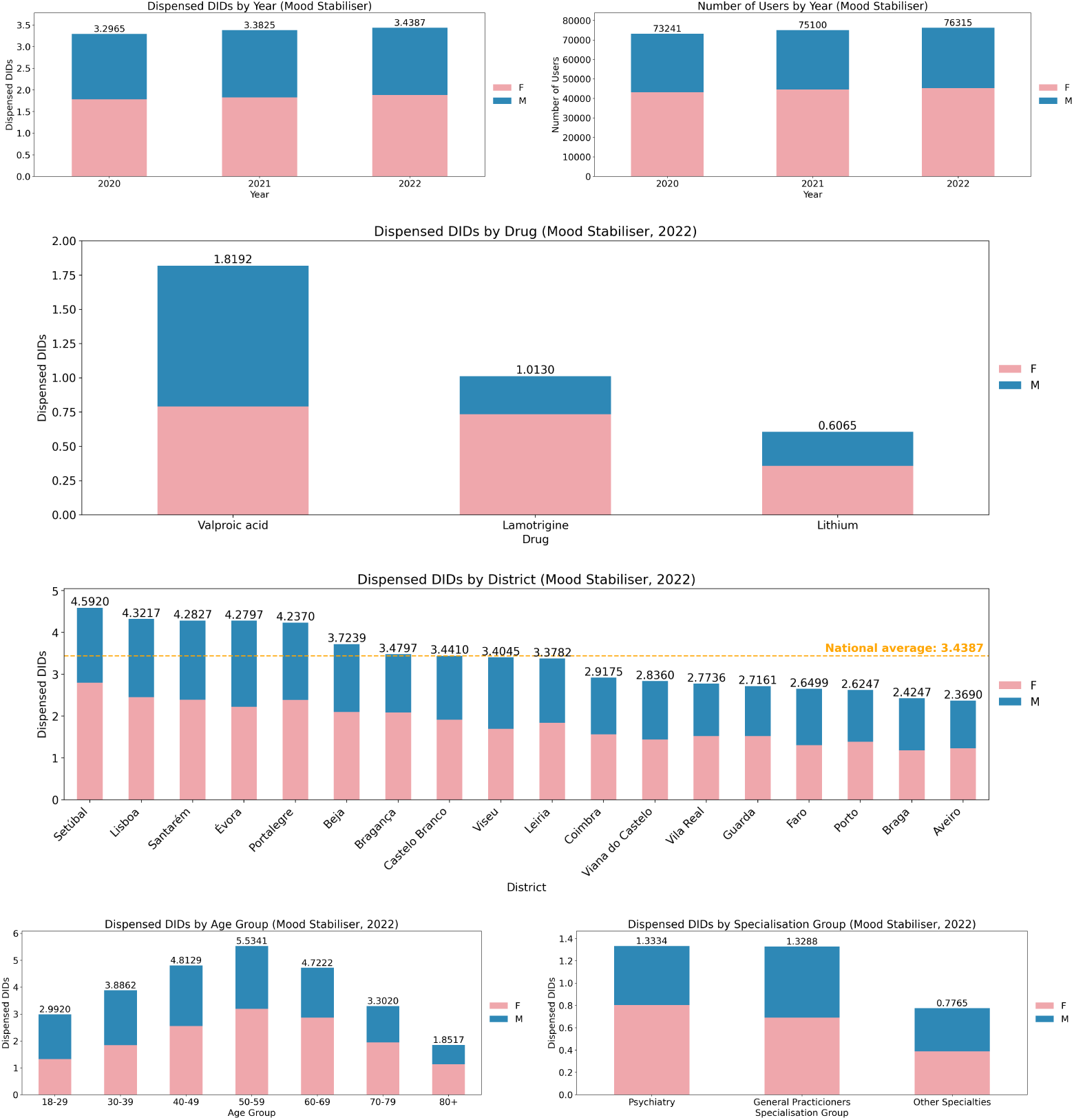
Mood stabiliser dispensation statistics (DIDs and user count) across districts, demographic sectors and medical specialties (gender is presented using stacked representations).

**Fig. 5.**
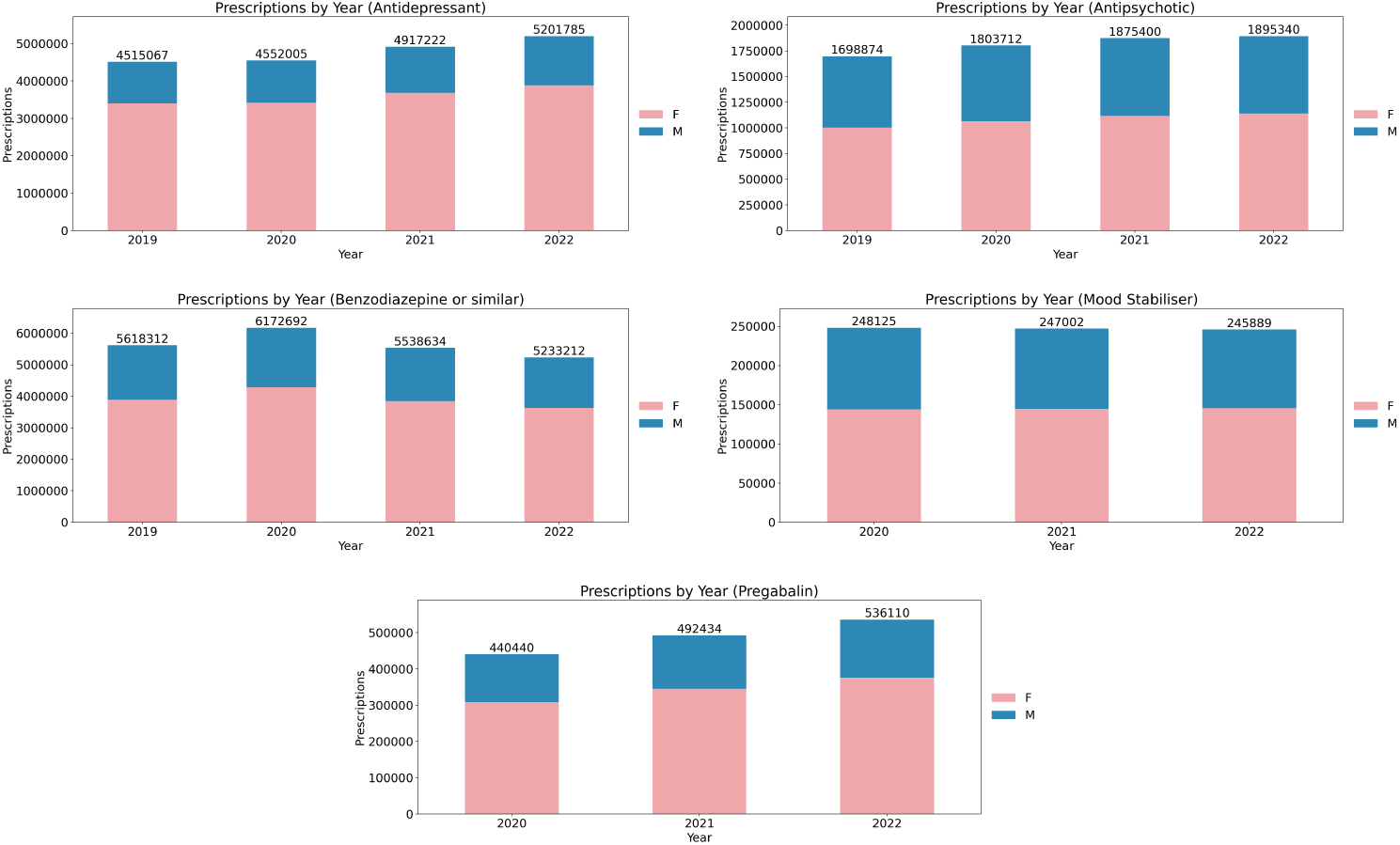
Evolution of the number of prescriptions with at least one dispensation act of each considered psychotropic drug class between 2019 (or 2020, if unavailable) and 2022.

**Fig. 6.**
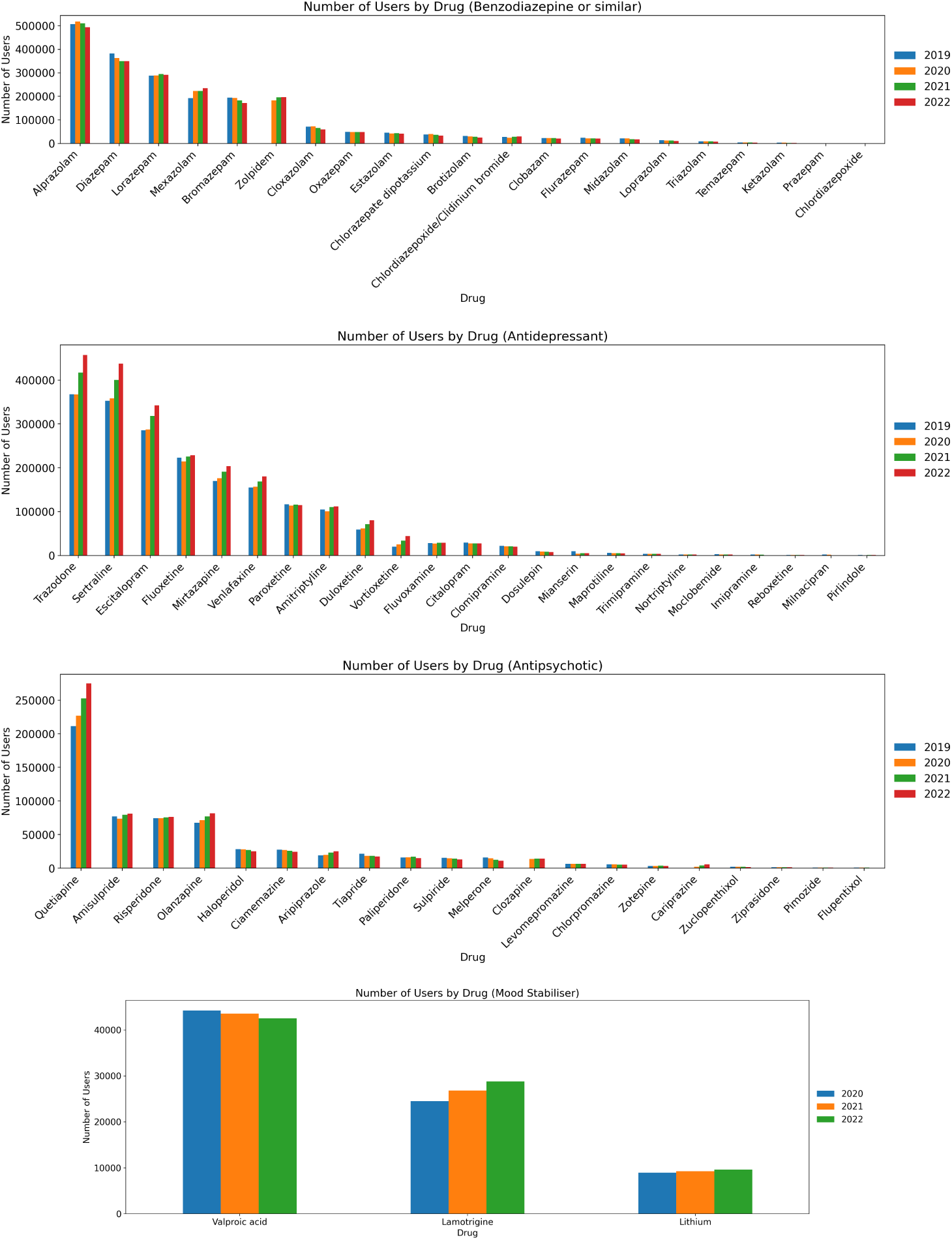
Number of users with at least one dispensation per psychotropic drug, grouped by their respective class, between 2019 (2020 for mood stabilisers) and 2022.

### 4.1 Benzodiazepines and zolpidem

Both the number of users (Figure 1) and the amount of dispensed DID (Table 1) in the benzodiazepine class has remained fairly stable – plateauing around the 1.7M users mark, over 15% of the Portuguese population. When looking at individual demographic groups, we can find some statistically significant growth trends, mainly driven by the male demographic, the 70-79 age group, and, in particular, the 18-29 age group, where we can find yearly DID growth trends in females and males of 9.59% and 7.47% of the 2019 baseline, respectively. In a more drug-centred analysis, the prescription of mexazolam has continued the growth trend shown since 2016, while bromazepam, on the contrary, has faced a relevant decrease.

In spite of these facts, the number of dispensed prescriptions of benzodiazepines has rapidly declined since 2020 (Figure 5).

### 4.2 Antidepressants

Regarding the antidepressant class, the growth trend exhibited since 2016 significantly accelerated in 2021 and 2022, suggesting an expected impact of the COVID-19 pandemic on the mental health of the Portuguese population, and, therefore, the consumption of drugs belonging to this class; as a result, it has replaced the benzodiazepine class as the most consumed psychiatric drug class by number of users, rapidly surpassing the 1.7M mark (Figure 2). This trend is also reflected in the majority of considered drugs and in all demographic classes, with emphasis on the 18-29 age group, where the average growth relative to the 2019 baseline (calculated from the regression coefficient) is above 15% per year in both males and females (Table 2).

### 4.3 Antipsychotics

In spite of the statistically significant growth trend since 2016 [27], the prescription and consumption of antipsychotics, fuelled by the majority of the age groups starting at 60 years old, along with the 18-29 age group in females, shows signs of slowing down, as shown by the statistically insignificant growth since 2019 (Table 3).

In particular, the number of users of quetiapine, risperidone, olanzapine, aripiprazole and cariprazine has significantly increased, as opposed to the remaining considered drugs, where the trend has remained stable or even decreased (Figure 3).

### 4.4 Mood stabilisers

Some demographic sections, mostly female, show statistically significant growth trends; however, the overall total falls below the 5% significance level (Table 4). Both the volume of expenditures and the State-funded copayment in this drug class were significantly impacted during this period by lithium, as a result of its global price growth registered between 2021 and 2022; with the drop registered in 2023 to prices similar to those in the beginning of 2021, it is expected to be reflected in the national expenditures in the near future (Figure 4).

An individualised analysis to lithium shows a slight but statistically significant consumption increase, which is particularly evident among older males (60 years and older) and in most female groups. When comparing age groups, a steep decline is observed in groups aged 70 and older.

The consumption metrics of valproic acid and lamotrigine must take in consideration their usage as antiepileptics, which may affect the analysis of this class under a psychiatry-centred lens.

## 5 DISCUSSION AND FINAL REMARKS

### 5.1 Benzodiazepines and zolpidem

Our results suggest a stable demand and prescription rate for this class of drugs in the general population. The high level of benzodiazepine use (over 15% of the population) raises questions about abuse, dependence and misuse, showing the need for careful monitoring [32] and development of guidelines to manage long-term use. Differences in trends in specific groups may indicate different levels of stress, changes in lifestyle or increased recognition and treatment of anxiety disorders in these specific demographic groups [26].

Table 1 suggests that significant increases appear to be specific to younger (as a result of anxiety and various adjustment reactions) [13] and older age groups (related to insomnia and behavioural disturbances) [1], suggesting a need for targeted public health interventions. Figure 1 supports several points:

(1) The consistent pattern of higher benzodiazepine use among women across years, districts and age groups seems to reflect differences in healthcare use;
(2) Some districts have higher DID than the national average, including Coimbra, Porto and Viseu, which supports the need for further research into overuse and development of targeted public health interventions;
(3) The use of benzodiazepines across districts could be partially explained by the distribution of psychiatrists across the country (with well known concentration of mental health professionals in the North of Portugal), and yet must be further explored, as Lisbon is, in this respect, “an outlier” – a place where there are well developed mental health services, but low prescription of benzodiazepines;
(4) GPs play a major role in benzodiazepine prescription, highlighting the importance of training and education policies at the primary care level for managing mental health conditions [34].

While diazepam continues to play a role far beyond psychiatry, justifying its use, alprazolam, lorazepam and mexazolam together gather for their well-known use in panic attacks, insomnia and generalised anxiety disorder [2, 36]. The increase in the consumption of mexazolam since 2016 [27] appears to be due to changing prescribing habits among healthcare professionals, particularly its very long half-life, low psychological and physical discontinuation effects, and safety [10]. The decrease in the number of prescriptions may indicate a shift towards prescribing larger quantities/boxes or changes in prescription guidelines affecting how often benzodiazepines can be prescribed.

In the case of zolpidem, despite the slight increase in its use, it was not noticeable or consistent across different age and gender categories. The exception is among men aged 30-39 years, which, together with the increased male use of benzodiazepines, could reflect a reduction of stigma towards mental health conditions or indicate underlying issues such as increased stress, anxiety or sleep problems in this particular age group.

### 5.2 Antidepressants

Our results show a marked increase in the use of antidepressants since 2016 [27], further accelerated in 2021 and 2022. This acceleration could be partly driven by the impact of the COVID-19 pandemic on mental health and the ensuing adjustment reactions with anxiety and depression. The increased consumption of antidepressants has made them the most prescribed class of psychotropic drugs in Portugal, indicating a significant change in mental health treatment patterns in Portugal and reflecting a clearer understanding of the need to address depression and anxiety disorders in a more treatment-oriented intervention rather than a palliative intervention (using benzodiazepines, for example) [8, 19].

While this increase was widespread across all age groups and genders, it particularly affected young adults (18-29 years), who experienced a 15% year-on-year increase compared to the 2019 baseline, highlighting how young adults may be experiencing stressors that drive adjustment responses, such as isolation, economic insecurity and stress. Table 2 shows that women are prescribed more antidepressants than men, which could be due to gender differences in the prevalence of depression [14], health behaviours or prescribing practices. Another explanation could be that stigma of mental illness might be limiting access of young men to psychotherapy and psychiatric care. It also shows higher use in older age groups, most likely reflecting the mental health impact of medical comorbidities, isolation or other age-related stressors. Finally, it shows that GPs are at the forefront of prescribing these medications and that there is a need for comprehensive training and advice in primary care.

Though rarely used as an antidepressant, trazodone occupies the forefront of antidepressant use in Portugal, showing how an off-label use of a drug can be so relevant – its use in the treatment of insomnia [18, 29]. Sertraline’s effectiveness, safety (including in pregnant women) and extensive approval for anxious and affective disorders and trauma makes the drug very attractive as the first line of treatment by GPs. Escitalopram and fluoxetine are two mature antidepressants that share a large market. Venlafaxine is the most prescribed SNRI and seems to be prescribed in more severe depressive episodes with psychomotor inhibition, or as a second line treatment after failing attempt with SSRI. The growth of vortioxetine, a recently introduced antidepressant in Portugal (2013 [15]), may come as a result of its specific profile in addressing cognitive symptoms in depression, as well as its lack of sexual side effects (which are the second reason – after weight gain – for maladaptation to therapy and dropouts).

The present findings may imply an increase in mental health awareness and education (reducing stigma and improving access to care), but also an increase in adjustment reactions or a lack of early access to counselling and therapy alongside pharmacotherapy. Ongoing research is needed to assess the long-term effects of increased use of antidepressants, including studies of their efficacy, safety and outcomes in different demographic groups.

### 5.3 Antipsychotics

The statistically significant increase in prescription and use of antipsychotics since 2016 [27] may suggest an increased recognition and treatment of psychotic disorders and bipolar disorder. However, considering that this is more prevalent in specific groups, we hypothesise this might be due to non-psychotic and off-label uses, such as the control of behavioural symptoms in dementia [40] and delirium [24] (e.g. in patients aged 60 and over), or the control of behavioural and anxiety symptoms in adjustment reactions or personality disorders (e.g. in women aged 18 to 29). Figure 3 supports that, similarly to the trends in other psychiatric medications, antipsychotics are more commonly prescribed to females and usage increases with age. Quetiapine and olanzapine are among the most dispensed antipsychotics, which may be due to their broad application (quetiapine in anxiety [28] and off-label use for insomnia [20] and bipolar disorder [21]; olanzapine for bipolar disorder [6], behavioural control in emergency departments and delirium [35]). However, the slow growth suggests that there may be a saturation point or a more conservative use due to concerns about side effects.

The reduction in expenditure appears to be due to price reductions – resulting from patent expiry, generic entry and renegotiation – of some active substances, namely aripiprazole, paliperidone, risperidone and quetiapine [15]. Long-term medication can be a financial burden, and cost management in health care ensures that treatments remain accessible and sustainable, allowing investment in innovation with newer treatments.

### 5.4 Mood stabilisers

The use of mood stabilisers has increased over the period of analysis in some demographic groups, suggesting an increase in the recognition or diagnosis of conditions requiring mood stabilisation and associated changes in clinical guidelines [25, 30], or possibly the stress of recent global events such as the COVID-19 pandemic. It cannot be excluded that the use of these drugs in neurology (changes in epilepsy management guidelines) may also be responsible for the increase.

A slight but statistically significant increase in lithium use was reported. This trend is particularly evident among older males (60 years and older) and in most female groups. The increase in older men is paradoxical given that most guidelines suggest that lithium should be avoided after the age of 65 because of its renal complications [33]. However, our results show a steep decline in the use of lithium over the age of 70, together with an increase in antipsychotics, supporting the changes in prescribing patterns in this age group. The increase in lithium in women may be due to increased awareness of good practice in mood stabilisation in women, where the alternative (valproic acid) is known to have teratogenic effects [37].

The large variation in lithium prices due to global price fluctuations could affect the access to this drug and disrupt the management of severe psychiatric conditions; therefore, policy makers and health care providers need to consider mechanisms to buffer against such price volatility.

### 5.5 General trends

Geographically, there is no clear information pointing towards a cross-sectional bias in the consumption of psychiatric drugs at the district level, although districts such as Coimbra and Viseu rank above the national average in consumption-related metrics in the three most consumed classes (antidepressants, antipsychotics and benzodiazepines), while Évora, Beja and Faro consistently rank below it. At a regional level, results indicate a higher consumption in the northern and central districts, in particular in the antidepressant and benzodiazepine classes.

The observed age distributions can be grouped into two main categories: i) antidepressants, benzodiazepines and zolpidem are skewed towards the older age groups when compared to the national distribution (over twice as much, in the case of the 80+ group), raising concerns over their overprescription; in contrast, ii) antipsychotics and mood stabilisers share similar curves that peak in the 50-59 group, with an additional, particularly high peak in the 80+ age group in the case of antipsychotics.

Regarding the prescriber’s specialty, the prevalences shown in the 2016-2019 period [27] remain generally true, with antidepressants and benzodiazepines (as well as zolpidem) being majorly prescribed by GPs (≥ 60%), and antipsychotics being prescribed by psychiatrists as the main specialty (44%), closely followed by GPs (40%). Additionally, while lithium is typically prescribed by psychiatrists, other considered mood stabilisers are mostly prescribed by GPs, likely as a result of their alternative use cases.

## Data Availability

All data produced in the present study are available upon reasonable request to the authors or to Serviços Partilhados do Ministério da Saúde.

## ACKNOWLEDGMENTS

Ethical approval was given by the Ethics Committee of CAML (*Centro Académico de Medicina de Lisboa*) with the reference number 340/20. The authors thank *Serviços Partilhados do Ministério da Saúde* (SPMS) for the data provision and valuable support, as well as *Guilherme Queiroz* for his precursor inputs. This work is further supported by national funds through *Fundação para a Ciência e Tecnologia* (FCT) under BII grant 2023/433 (PTDC/CCI-CIF/4613/2020) to D.B. and the INESC-ID pluriannual (UIDB/50021/2020).

## A PREGABALIN RESULTS AND DISCUSSION

An increase in the consumption of pregabalin (dispensed DIDs) has been registered by the majority of demographic segments (predominantly female), a trend followed by the number of active users, as Table 5 and Figure 7 suggest. The sharp increase in its consumption is most likely driven not only for its anxiolytic properties, but by complementary uses in neurology as an antiepileptic [12] and in neuropathic pain as an analgesic [4]. The important role of GPs in prescribing mood stabilisers highlights the need for primary care providers to be well informed and up to date with mental health practices.

**Table 5.** Evolution of dispensed DIDs in pregabalin between 2019 and 2022.

| Gender | Age group | 2020 | 2021 | 2022 | Mean change per year, fraction of baseline | P | 95% confidence interval | r<sup>2</sup> |
| --- | --- | --- | --- | --- | --- | --- | --- | --- |
| F | 18-29 | 0.0311 | 0.0358 | 0.0426 | 0.1860 | 0.0677 | [-0.0662, 0.4382] | 0.9887 |
|  | 30-39 | 0.0805 | 0.0910 | 0.0966 | 0.1001 | 0.1128 | [-0.1276, 0.3278] | 0.9689 |
|  | 40-49 | 0.2756 | 0.3040 | 0.3317 | 0.1016 | 0.0040 | [0.0934, 0.1098] | 1.0000 |
|  | 50-59 | 0.5194 | 0.5736 | 0.6225 | 0.0993 | 0.0189 | [0.0619, 0.1367] | 0.9991 |
|  | 60-69 | 0.6721 | 0.7417 | 0.8190 | 0.1092 | 0.0191 | [0.0677, 0.1507] | 0.9991 |
|  | 70-79 | 0.6487 | 0.7256 | 0.8103 | 0.1245 | 0.0180 | [0.0797, 0.1693] | 0.9992 |
|  | 80+ | 0.4656 | 0.5079 | 0.5603 | 0.1018 | 0.0390 | [0.0224, 0.1812] | 0.9962 |
|  | Total | 2.6931 | 2.9796 | 3.2830 | 0.1095 | 0.0105 | [0.0866, 0.1324] | 0.9997 |
| M | 18-29 | 0.0228 | 0.0239 | 0.0303 | 0.1653 | 0.2438 | [-0.6808, 1.0114] | 0.8603 |
|  | 30-39 | 0.0576 | 0.0631 | 0.0695 | 0.1035 | 0.0236 | [0.0548, 0.1522] | 0.9986 |
|  | 40-49 | 0.1335 | 0.1504 | 0.1593 | 0.0968 | 0.1123 | [-0.1225, 0.3161] | 0.9692 |
|  | 50-59 | 0.2140 | 0.2391 | 0.2654 | 0.1200 | 0.0090 | [0.0984, 0.1416] | 0.9998 |
|  | 60-69 | 0.3031 | 0.3366 | 0.3665 | 0.1046 | 0.0207 | [0.0614, 0.1478] | 0.9989 |
|  | 70-79 | 0.3088 | 0.3349 | 0.3723 | 0.1027 | 0.0657 | [-0.0325, 0.2379] | 0.9894 |
|  | 80+ | 0.1879 | 0.2067 | 0.2281 | 0.1070 | 0.0238 | [0.0562, 0.1578] | 0.9986 |
|  | Total | 1.2277 | 1.3547 | 1.4914 | 0.1074 | 0.0135 | [0.0783, 0.1365] | 0.9995 |
| Total |  | 3.9208 | 4.3343 | 4.7744 | 0.1089 | 0.0114 | [0.0840, 0.1338] | 0.9997 |

**Fig. 7.**
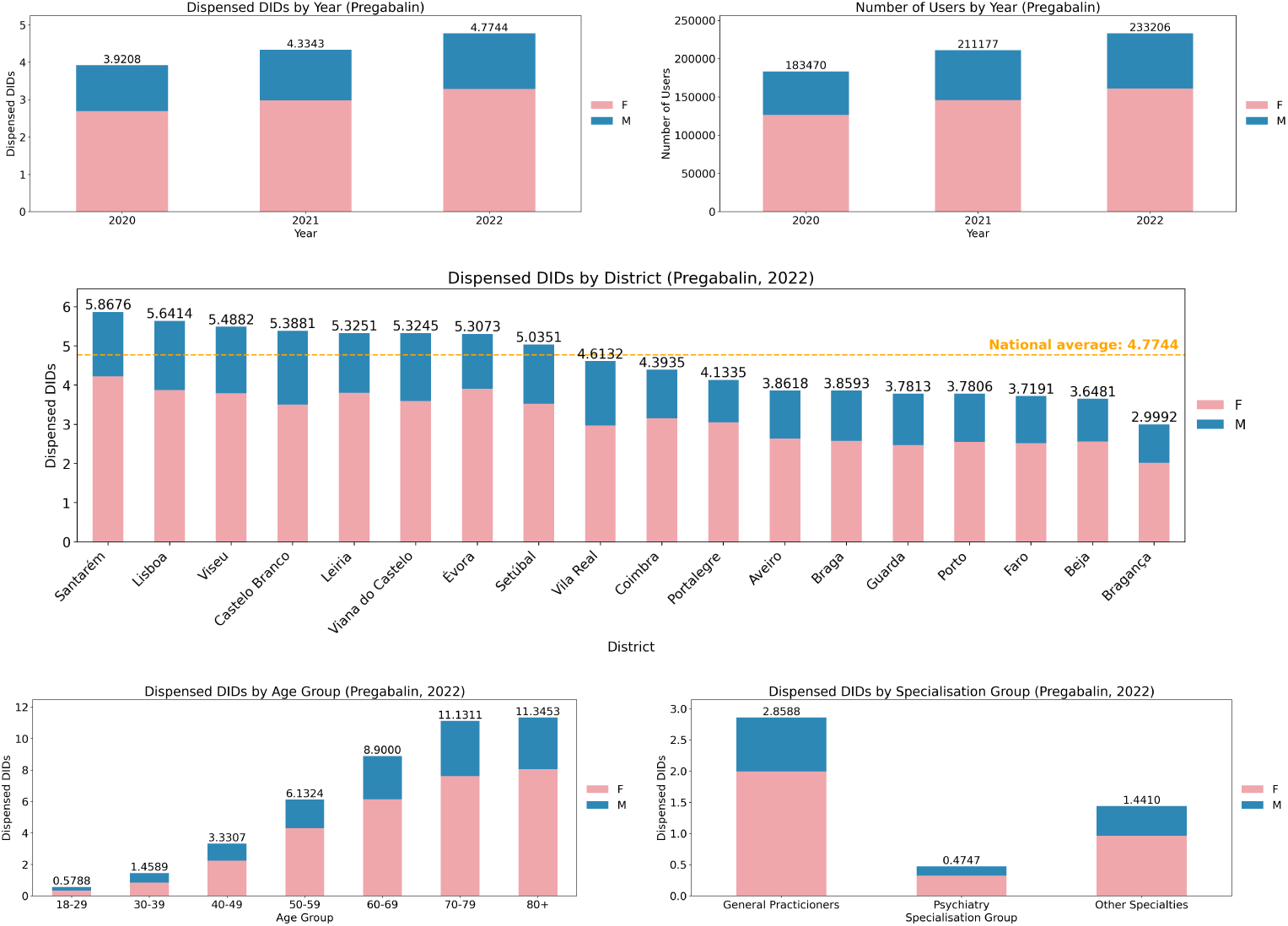
Pregabalin dispensation statistics (DIDs and user count) across districts, demographic sectors and medical specialties (gender is presented using stacked representations).

## B SUPPLEMENTARY RESULTS

**Fig. 8.**
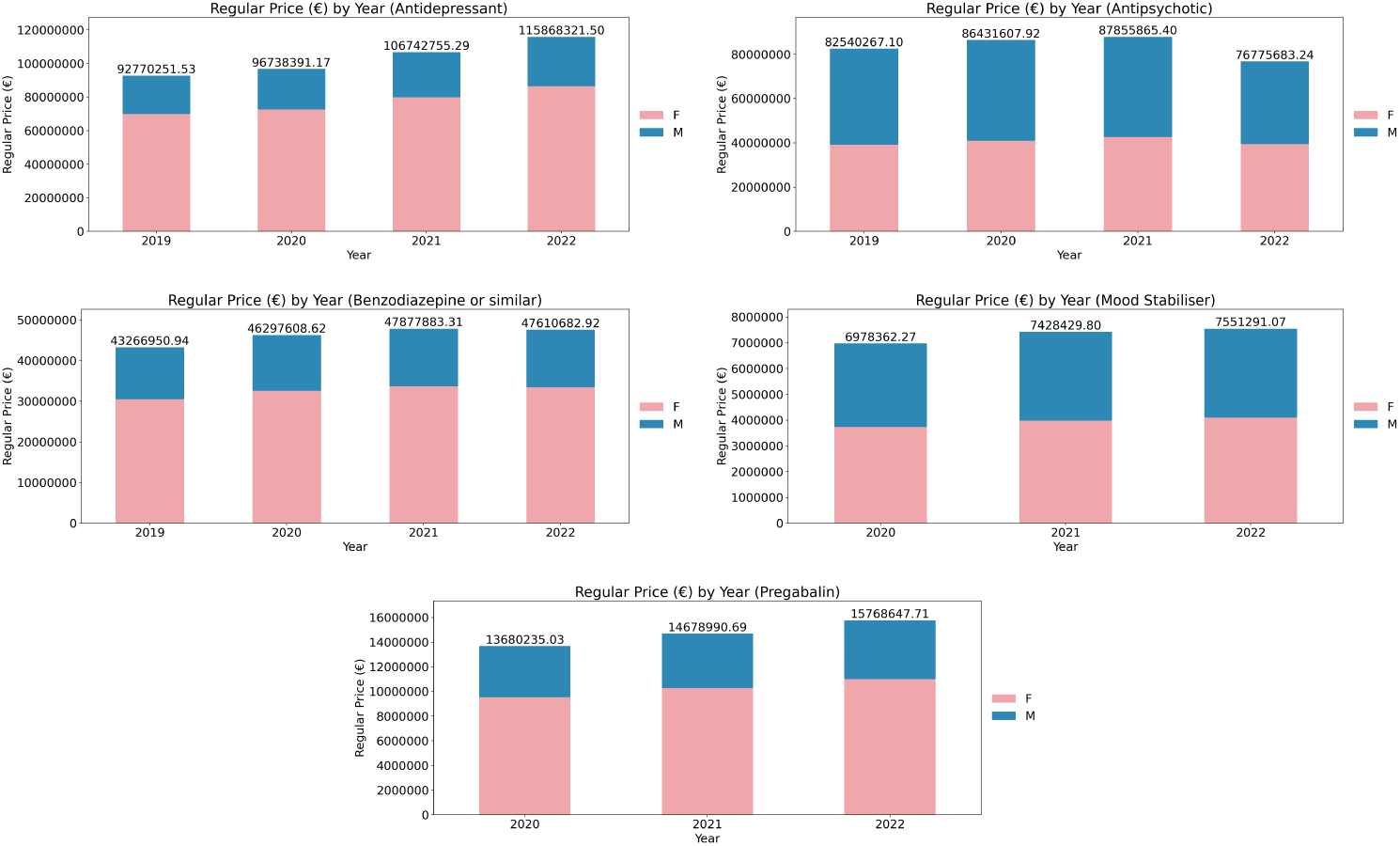
Evolution of the total expenditure of each considered psychotropic drug class between 2019 (or 2020, if unavailable) and 2022.

**Fig. 9.**
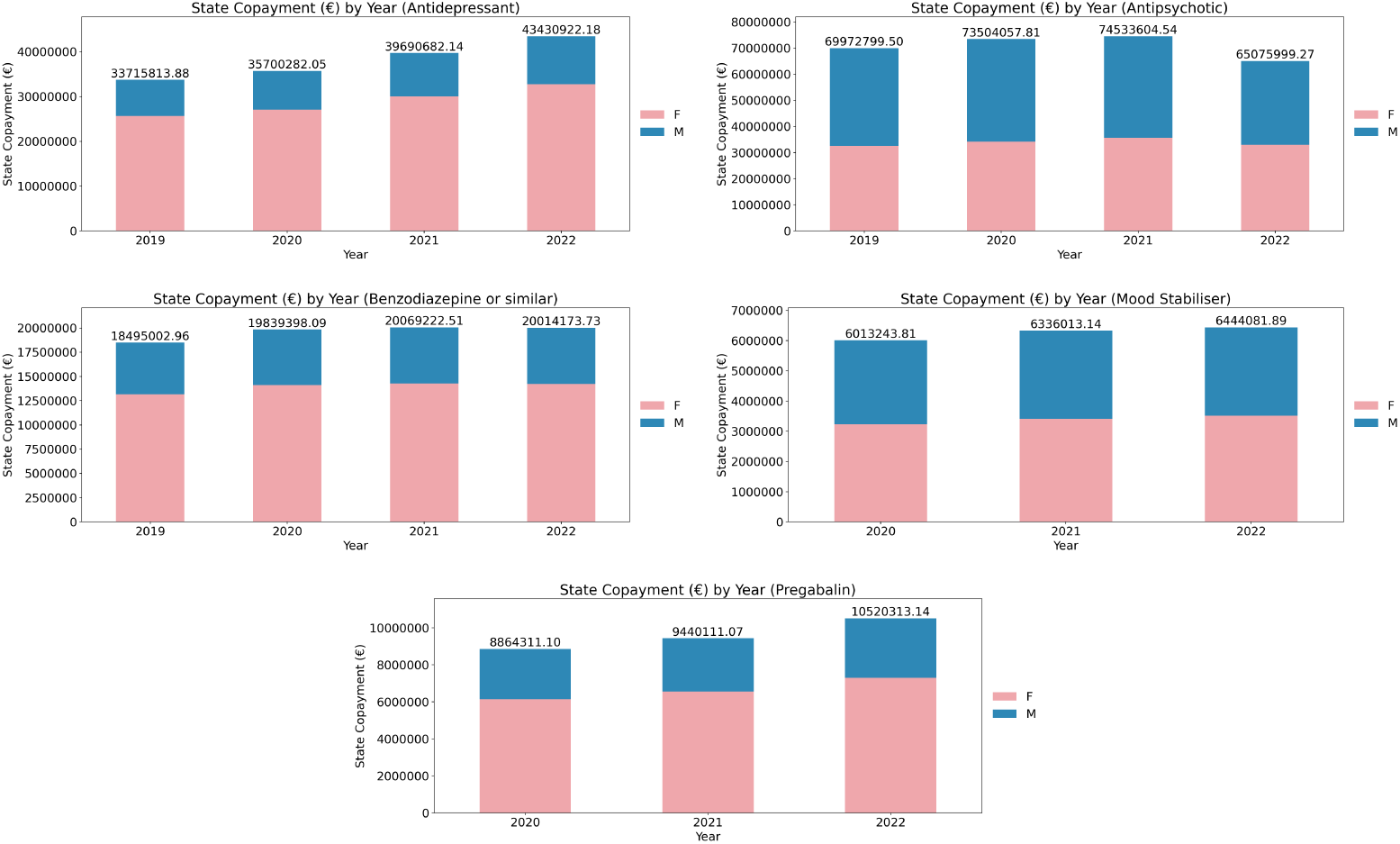
Evolution of the total state-funded copayment of each considered psychotropic drug class between 2019 (or 2020, if unavailable) and 2022.

## C LIST OF AVAILABLE DRUG PACKAGES

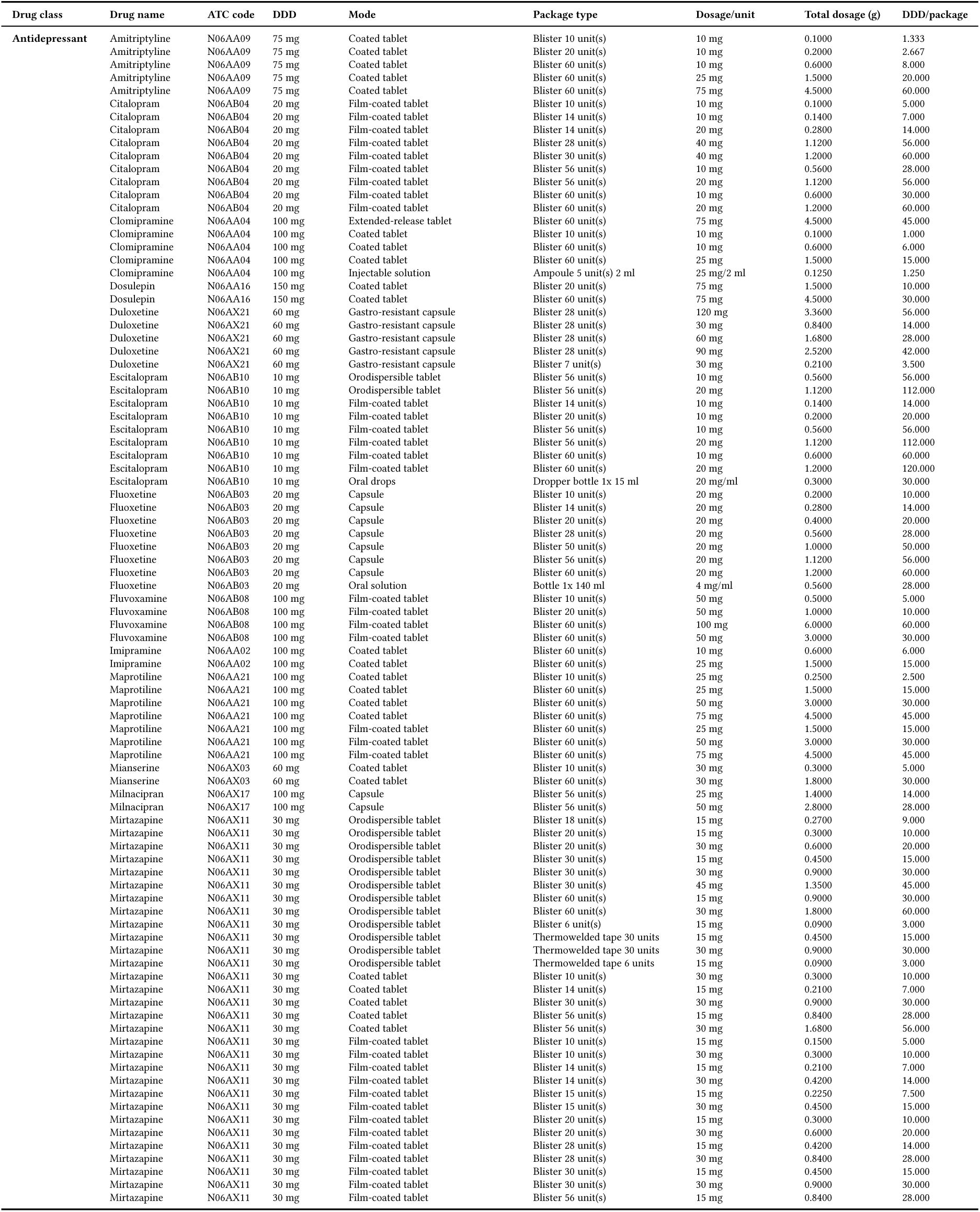
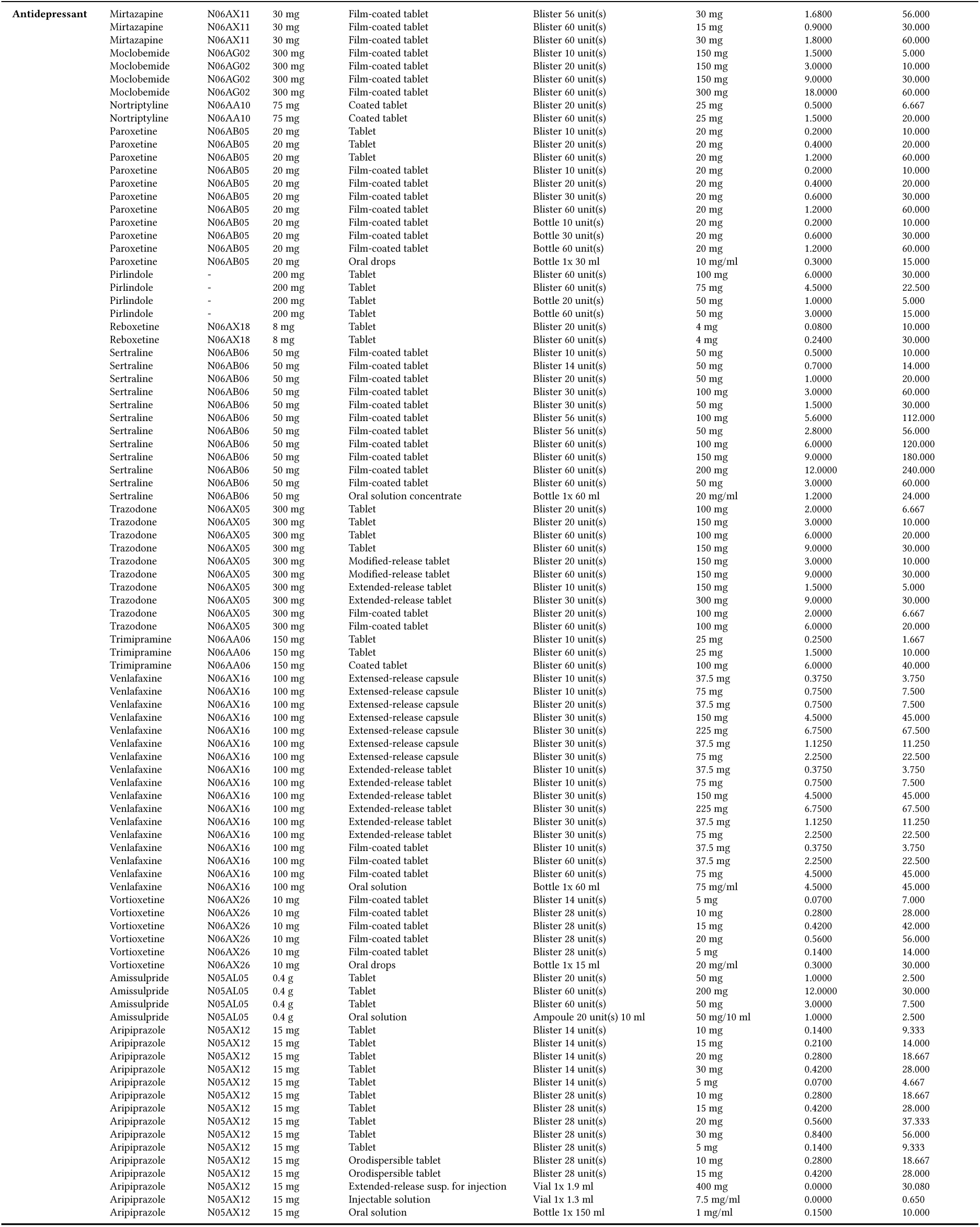
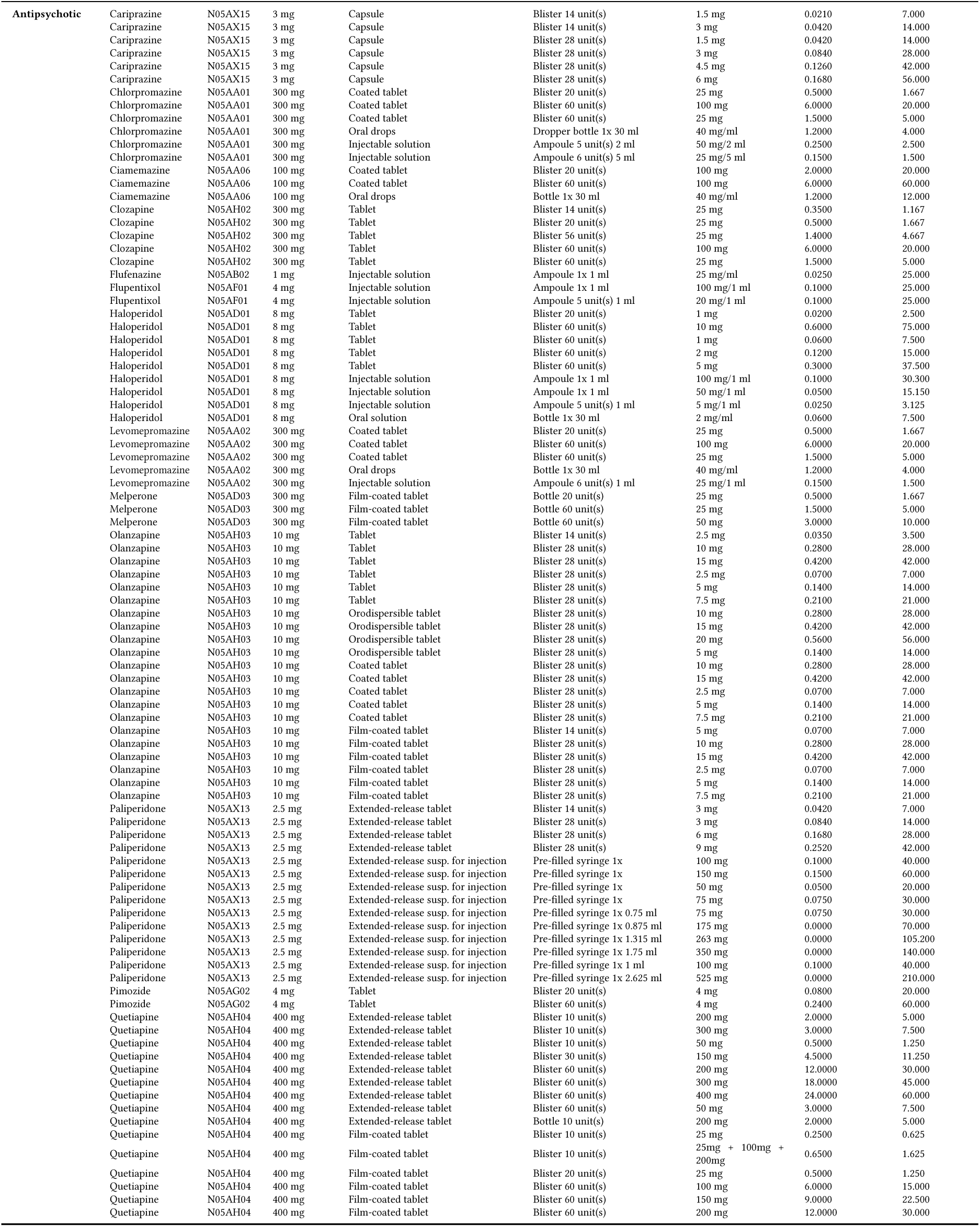
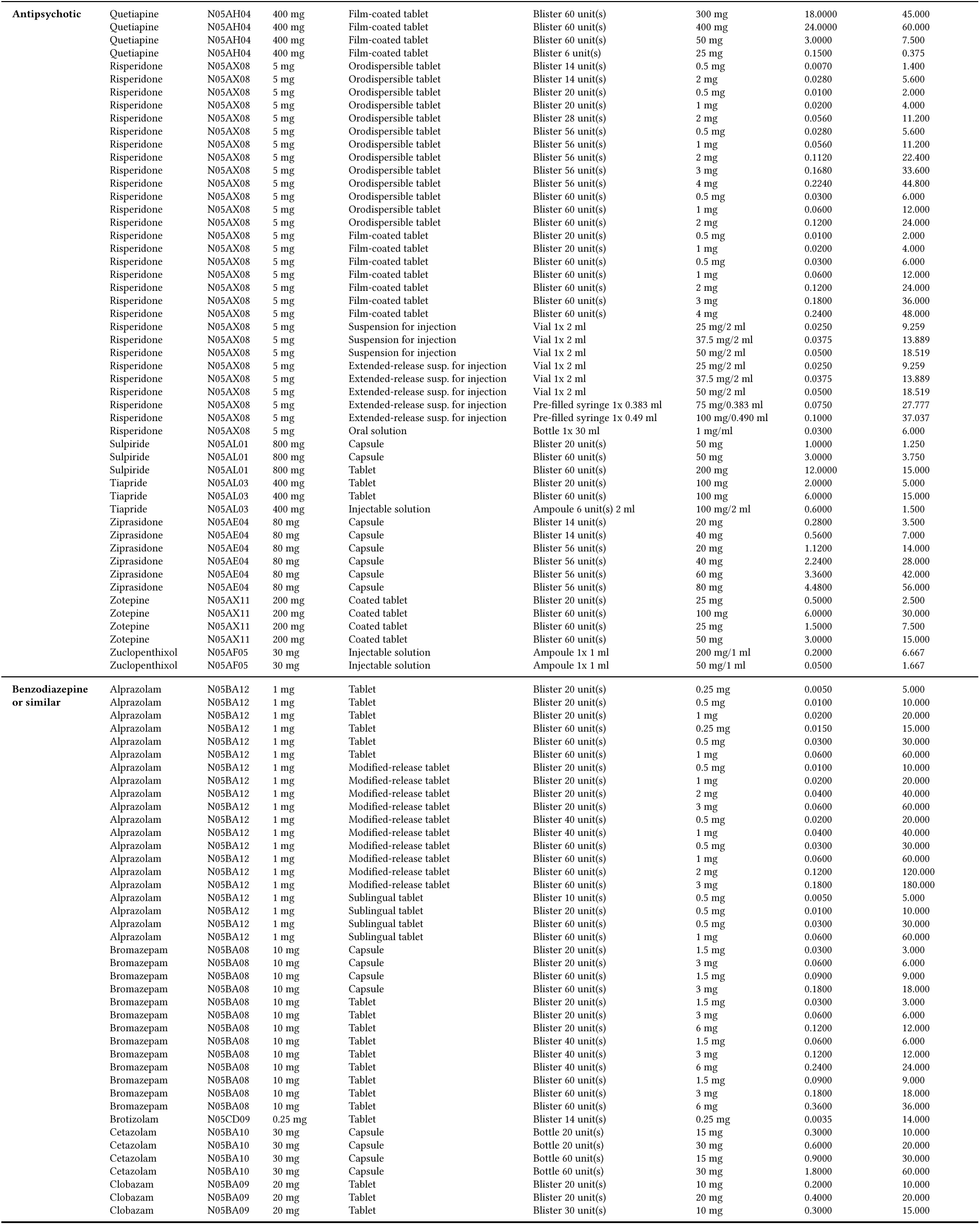
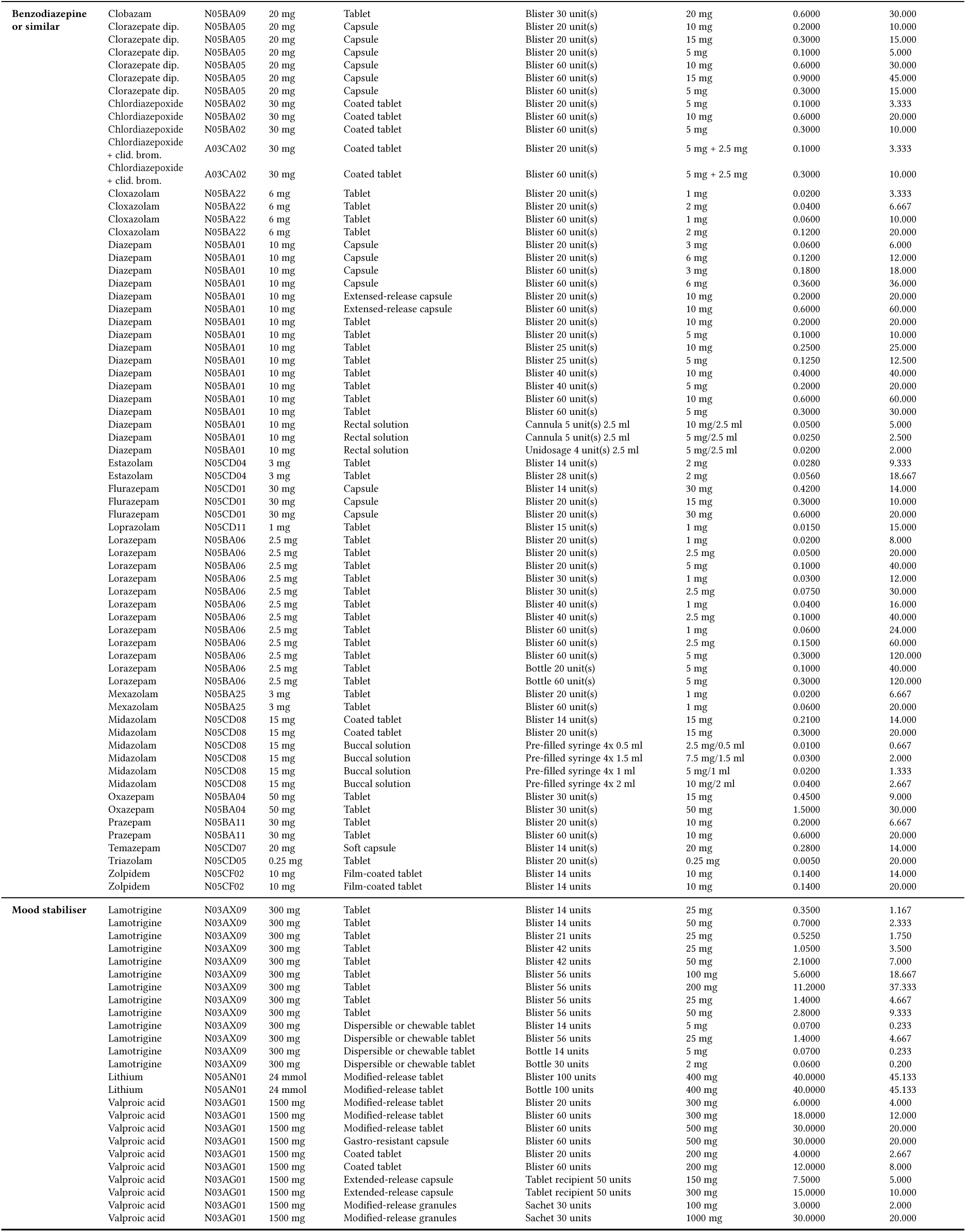
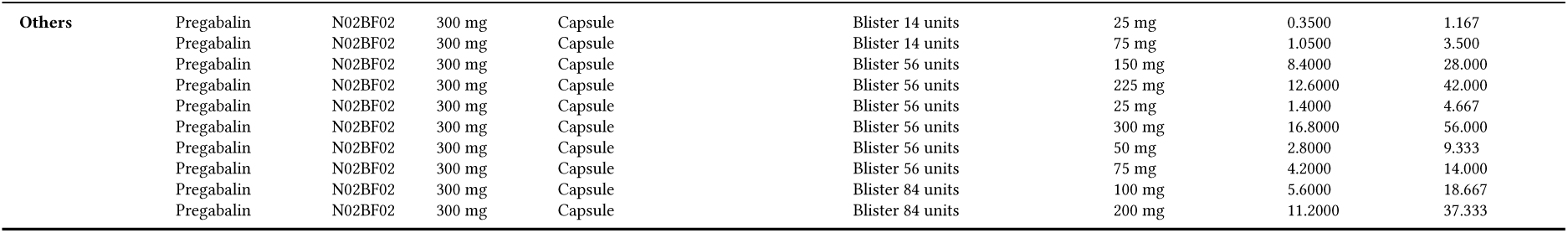

## Footnotes

1 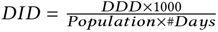, where a DDD (Defined Daily Dose) is defined by WHO as the average maintenance dose per day for a drug used for its main indication in adults [41].

